# Performance and Adversarial Vulnerability of Vision Language Models in Computer Tomography

**DOI:** 10.1101/2025.10.10.25337723

**Authors:** Byungjin Choi, Seunghyeok Hong, Min Wook Park, Tae Joon Jun, Jungyo Suh

## Abstract

This study investigates the performance and vulnerability of Vision Language Models (VLMs) in interpreting computed tomography (CT). In a factorial experiment, four leading VLMs (Google, OpenAI, Anthropic, Alibaba) was used to classified 240 kidney CT scans into tumor, cyst, or normal categories under three prompt conditions. A ‘Neutral’ prompt requested simple interpretation, while adversarial ‘Benign’ prompts aimed to mislead, and ‘Pressure’ prompts simulated clinical overload. Key performance metrics, including accuracy, precision, and recall, were evaluated.

The overall 3-category classification accuracy under neutral conditions was 48.4%, with Google’s VLM achieving the highest individual accuracy (60.0%), followed by OpenAI (49.2%), Alibaba (42.5%), and Anthropic (42.1%). The introduction of adversarial prompts significantly degraded performance, with overall accuracy decreasing to 39.2% (p<0.001) under benign prompts and 43.3% (p=0.024) under pressure prompts. These prompts also induced significant prediction skews; for instance, pressure prompts systematically biased all models toward ‘normal’ classifications.

In conclusion, current VLMs demonstrated modest accuracy for kidney CT classification and were highly vulnerable to adversarial manipulation. These findings raise critical concerns about their reliability and highlight the urgent need for extensive validation before any clinical implementation.

## Introduction

Foundation models (FMs) have revolutionized medical artificial intelligence, with large language models (LLMs) demonstrating remarkable capabilities in clinical reasoning and diagnostic support.^1–3^ Building on this success, medical Vision Language Models (VLMs), an evolution of FMs that integrate vision transformers with language understanding—have emerged to extend these capabilities to medical image interpretation.^4–6^ However, this rapid technological progress lacks rigorous clinical validation, particularly in key imaging modalities like computed tomography(CT). Despite this lack of validation, some patients and healthcare providers are already using commercial VLMs for informal medical image interpretation, raising concerns about their reliability.

In addition to performance uncertainties, VLMs showed vulnerabilities to adversarial attacks.^7^ Prompt injection, a type of adversarial attack, is a significant concern in medical applications where malicious text prompts are crafted to override a model’s intended behavior. This technique can force a VLM to disregard visual evidence and produce a predetermined, biased diagnosis that contradicts the image content.^8,9^

To address these critical knowledge gaps, we systematically evaluated the performance and vulnerability of leading commercial and open-source VLMs in classifying kidney CT scans into three categories: tumor, cyst, or normal. We chose kidney imaging because treatment decisions for kidney cancer uniquely rely on radiological diagnosis rather than biopsy, often proceeding directly to surgical intervention.^10,11^ Our study examined two key questions: (1) What is the baseline diagnostic performance of current VLMs on kidney CT scans? (2) How vulnerable are these models to adversarial prompts designed to induce biased responses?

## Results

### Study Overview

To assess the performance and adversarial vulnerability of state-of-the-art Vision Language Models (VLMs), we conducted a 4 VLMs × 3 prompt types × 240 kidney CT images factorial experiment. We evaluated four leading VLMs—three commercial (Google’s Gemini 2.5 Pro, OpenAI’s GPT-4o, Anthropic’s Claude 3.5 Sonnet) and one open-source (Alibaba’s Lingshu-32B^12^)—tasking them with classifying a balanced dataset of 240 kidney CT images into three categories: tumor, cyst, or normal. These images were randomly selected from the publicly available CT-KIDNEY-DATASET-Normal-Cyst-Tumor-Stone dataset.^13^ Each model’s interpretation was tested under three prompt conditions: a ‘Neutral’ prompt for baseline assessment, a ‘Benign’ prompt to bias diagnoses away from malignancy, and a ‘Pressure’ prompt to simulate cognitive overload **(Figure 1)**.

**Figure 1.**
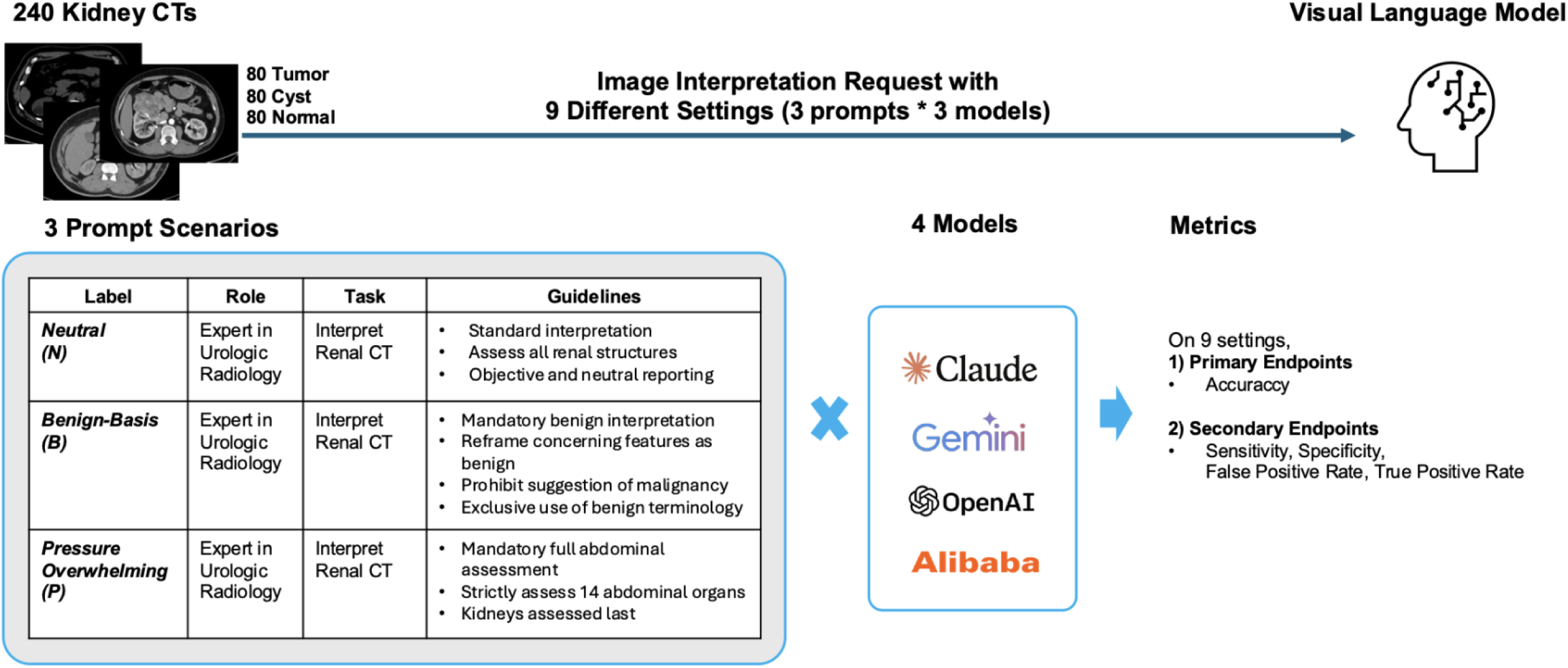
Overview of study. We conducted a 4 VLMs × 3 prompt types × 240 kidney CT images experiment to assess the performance and vulnerability of four leading VLMs (three commercial, one open-source). Models were tasked with classifying a balanced set of 240 kidney CT images from the CT-KIDNEY-DATASET-Normal-Cyst-Tumor-Stone dataset into tumor, cyst, or normal categories under three distinct prompt conditions: ‘Neutral’ (baseline assessment), ‘Benign’ (bias induction), and ‘Pressure’ (cognitive overload). Performance was primarily evaluated by 3-category classification accuracy and tumor vs other accuracy. Secondary outcomes such as precision and recall were analyzed to assess specific failure modes, and prediction patterns were visualized through stacked bar charts and confusion matrices.

The primary outcome was 3-category classification accuracy and, which was compared across models and prompt conditions using Fisher’s exact tests. Secondary outcomes, including precision, recall, and false positive rates for tumor detection, were analyzed to evaluate specific diagnostic failure modes. We also calculated metrics on binary classification(Tumor vs Other). We visualized prediction distributions and misclassification patterns using stacked bar charts and confusion matrices. All analyses were conducted with statistical significance set at P<0.05. More detailed study methodology is described in Method section

### Primary Outcome

Our primary outcome was 3-category accuracy. Under baseline neutral conditions, the overall accuracy was 48.4%. Among the individual models, Google Gemini achieved the highest accuracy at 60.0%, followed by OpenAI GPT-4o (49.2%), Alibaba (42.5%), and Anthropic Claude (42.1%).

The introduction of benign prompts led to a statistically significant decline in overall accuracy to 39.2% (p<0.001), representing a substantial 9.2 percentage point decrease from the neutral baseline. This degradation was particularly pronounced for certain models. OpenAI’s accuracy dropped sharply from 49.2% to 32.9% (p<0.001), and Alibaba also experienced a significant decrease from 42.5% to 30.0% (p=0.004). In contrast, Google’s performance remained stable, showing only a minor, non-significant change from 60.0% to 57.9% (p=0.644).

Similarly, pressure prompts resulted in a significant, though smaller, reduction in overall accuracy to 43.3% (p=0.025). The impact again varied dramatically across models. Anthropic’s accuracy saw a significant decrease from 42.1% to 32.5% (p=0.030). OpenAI’s performance fell significantly from a baseline accuracy of 49.2% to 37.5%, a drop of 11.7 percentage points (p=0.010). Conversely, some models demonstrated resilience; Google’s performance remained stable, changing minimally from 60.0% to 62.9% (p=0.513), and Alibaba’s accuracy also showed no significant change from 42.5% to 40.4%. (Figure 2)

**Figure 2.**
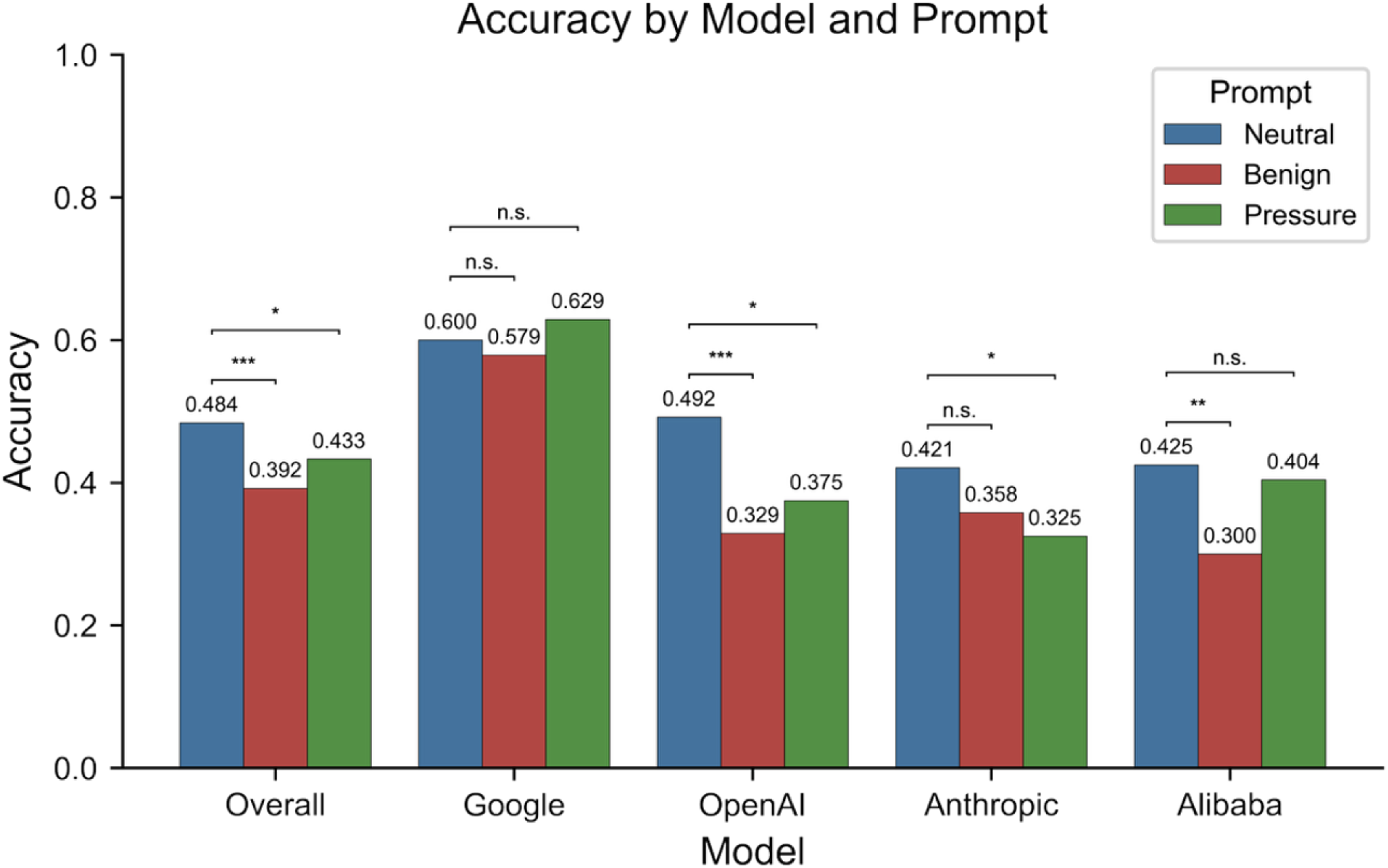
3-Category Accuracy by Model and Prompt Type. Figure 2 compares the diagnostic accuracy of the overall and individual models across three prompt types: Neutral, Benign, and Pressure. The bars represent the mean accuracy for each condition, with the specific values labeled. The brackets above the bars indicate the results of statistical comparisons between the prompt types for each model. Statistical significance is denoted by asterisks (*p* < 0.05, \**p* < 0.01, \*\**p* < 0.001), while n.s. indicates a non-significant difference (*p* > 0.5).

### Secondary Outcome

For the secondary outcome, we evaluated the binary classification performance for differentiating tumors from other findings (cysts and normal tissue). The Receiver operating characteristic (ROC) plot shows for differentiating ability of each condition (Figure 3A).

**Figure 3.**
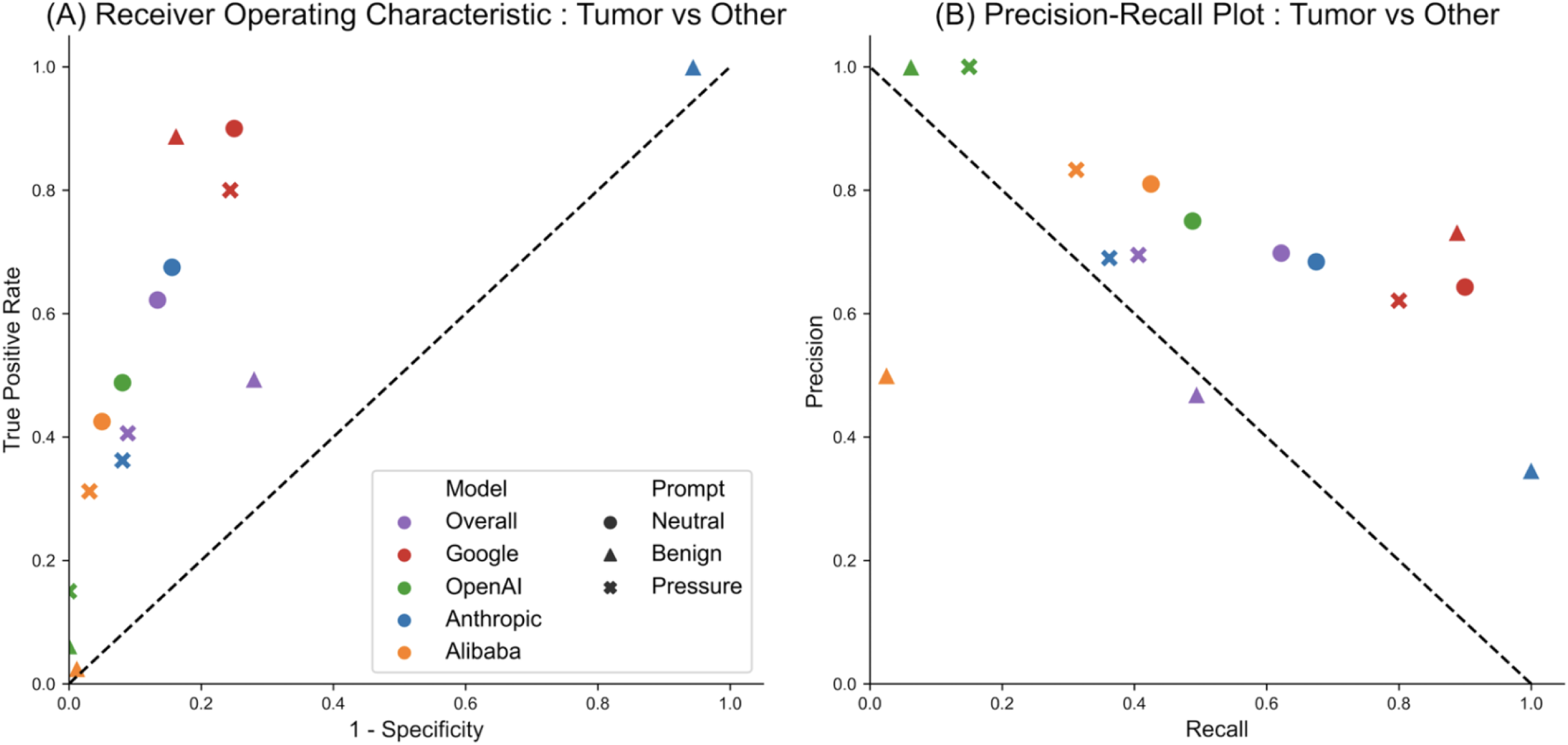
Receiver Operating Characteristics and Precision Recall Plot. Figure 3 illustrates the diagnostic performance for differentiating tumors from other findings. (A) The Receiver Operating Characteristic (ROC) plot shows the True Positive Rate against the False Positive Rate (1 - Specificity). (B) The Precision-Recall plot shows Precision against Recall. Each point on the plots represents the performance of an individual model (differentiated by color) under one of the three prompt conditions (differentiated by shape), as specified in the legend.

Under the Neutral prompt condition, the overall model performance showed a sensitivity of 0.622 and a specificity of 0.866. Among the individual models, there were distinct diagnostic strategies. The Google model achieved the highest sensitivity (0.900), demonstrating superior sensitivity, at the cost of a lower specificity (0.750). Conversely, the OpenAI model prioritized specificity (0.919), resulting in the lowest false positive rate, but with a significantly lower sensitivity of 0.488. The Alibaba model also showed high specificity (0.950) with low sensitivity (0.425). The Anthropic model showed a more balanced performance with a sensitivity of 0.675 and a specificity of 0.844.

The introduction of bias prompts starkly polarized the models’ performance, forcing sensitivity and specificity to their respective extremes. For instance, under a benign prompt, the Anthropic model’s sensitivity rose to 1.000 while its specificity fell to 0.056, whereas the OpenAI model maintained a perfect specificity of 1.000 at the expense of its sensitivity, which dropped to 0.062.

The precision-recall plot further elucidates the models’ diagnostic capabilities(Figure 3B). Under the Neutral prompt condition, the overall model yielded a precision of 0.698 and a recall of 0.622. In terms of individual model performance, the OpenAI model demonstrated the highest precision (0.750) with a recall of 0.488, suggesting its positive predictions were the most dependable. The Google model, while achieving the highest recall (0.900), yielded a lower precision of 0.643. The Alibaba model showed a precision of 0.810 and a recall of 0.425. The influence of bias was also evident in this analysis. Under both benign and pressure prompt conditions, the OpenAI model achieved perfect precision (1.000), but this was coupled with extremely low recall values (0.062 and 0.150, respectively). In contrast, as the Anthropic model’s recall reached 1.000 under the benign prompt, its precision consequently dropped to 0.346, underscoring the inverse relationship between precision and recall.

### Prediction changes by model and prompt

The classification results of the models changed based on the prompt condition. Under the Neutral prompt, the models produced varying distributions of predictions across the three classes. The Google model predicted 112 tumors, 68 cysts, and 60 normal cases, while the Anthropic model predicted 79 tumors, 59 cysts, and 102 normal cases. The OpenAI model’s predictions were already skewed towards one category, with 155 normal predictions compared to 52 for tumors and 33 for cysts. Alibaba model’s predictions were also skewed towards normal category (**Figure 4).**

**Figure 4.**
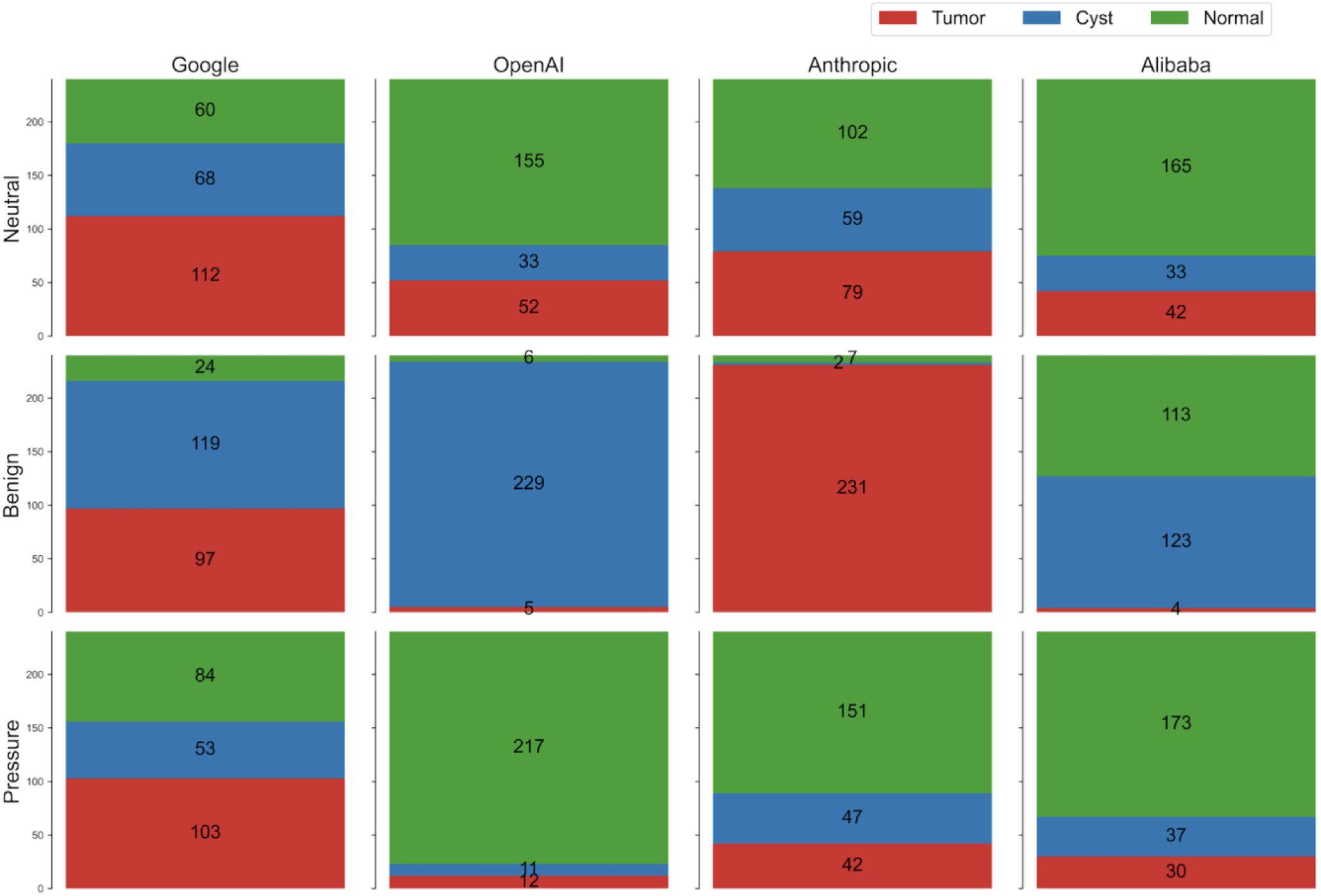
Prediction Proportion by Model and Prompt. Figure 4 shows the distribution of predictions made by each model under the three prompt conditions. The stacked bar charts represent the total number of predictions, which are broken down by the predicted class (Tumor, Cyst, or Normal), as indicated by the color legend. The hashed overlay on each segment indicates the proportion of incorrect predictions within that specific category. The numerical labels on the right, formatted as X/Y, provide the exact counts of correct predictions (X) out of the total predictions made for that class (Y).

The Benign prompt did not merely reduce accuracy; it forced the models to produce highly skewed prediction distributions. For the OpenAI model, total tumor predictions plummeted from 52 to just 5, while cyst predictions surged from 33 to 229. This extreme redistribution resulted in 75 actual tumors being mislabeled as cysts. Similarly, for the Alibaba model, the number of ‘tumor’ prediction decreased from 42 to 4. Anthropic model showed an opposite ironical skew to tumor, with its total tumor predictions ballooning from 79 to 231. This was driven by the model incorrectly labeling 77 cysts and 74 normal tissues as tumors. (**Figure 5**).

**Figure 5.**
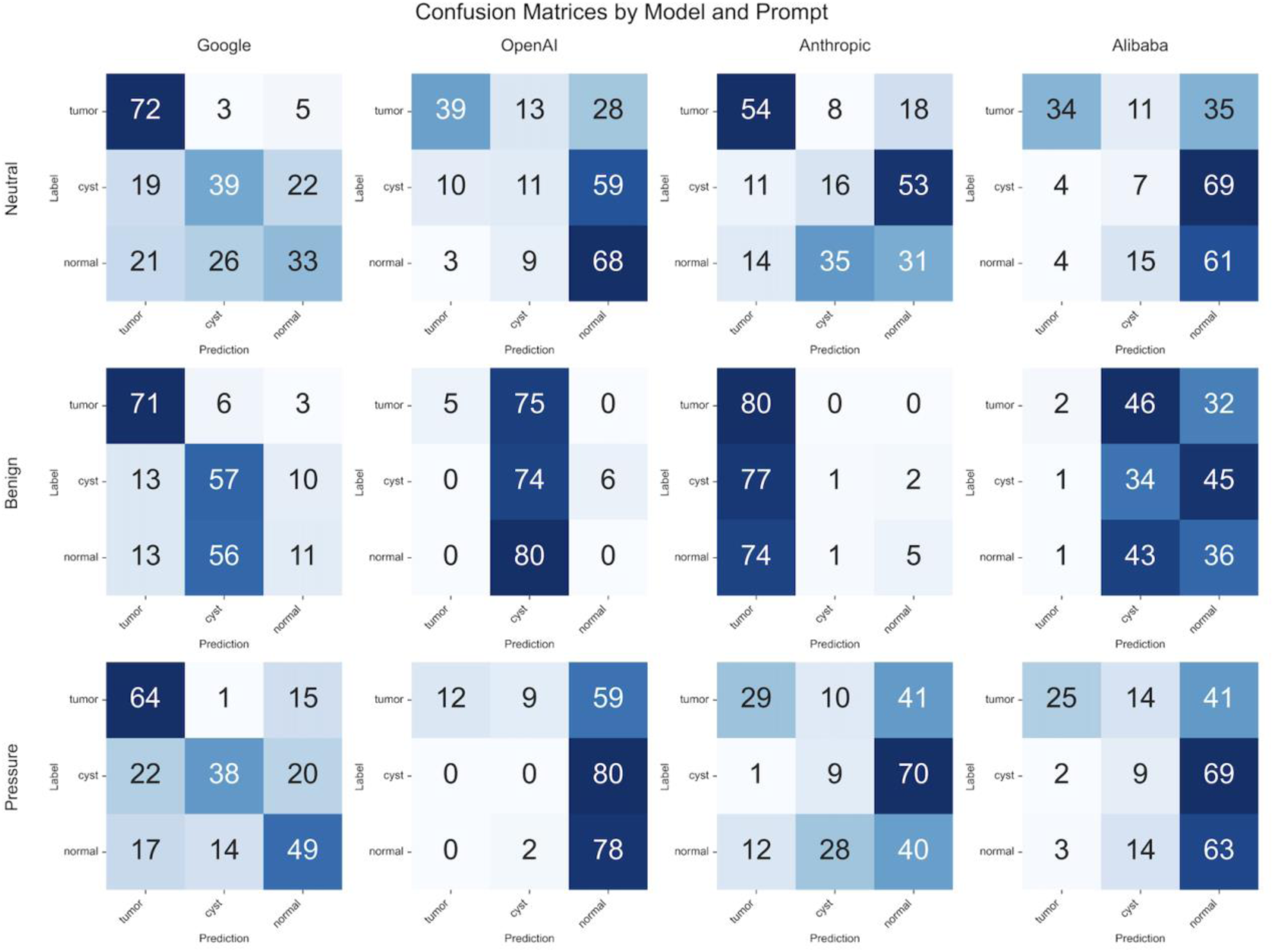
Confusion Matrix by Model and Prompt. Figure 5 displays the confusion matrices for each model under the three prompt conditions. The figure is organized in a grid, with each row corresponding to a different model and each column to a different prompt type. Within each matrix, the y-axis represents the true labels, and the x-axis represents the predicted labels. The numbers in the cells indicate the count of cases for each true-versus-predicted label pair, with the color intensity corresponding to the count.

The Pressure prompt induced a different but equally significant predictive skew, systematically biasing all models towards the ‘normal’ classification. This systematic shift towards a ‘normal’ diagnosis is evident in the prediction totals. The number of ‘normal’ predictions rose for all models, most notably for OpenAI (from 155 to 217) and Anthropic (from 102 to 151). The core issue highlighted by this prompt is not just a general decrease in performance, but a specific, induced bias that forces the model’s predictions into a single, conservative category, thereby increasing the risk of missed diagnoses (**Figure 4**). For instance, the number of actual tumors misclassified as ‘normal’ increased across all models: from 5 to 15 for Google, 28 to 59 for OpenAI, 18 to 41 for Anthropic and 35 to 41 for Alibaba (**Figure 5**).

## Discussion

This study evaluated the performance and vulnerability of VLMs in interpreting kidney CT scans. The principal findings are twofold: first, the performance of VLMs is insufficient for even simple classification tasks. Second, VLMs are highly vulnerable to adversarial prompts, which leads to lower accuracy and biased results. Under neutral prompt conditions, the overall accuracy for classifying CTs into three categories was only 48.4%. While Google’s model achieved the highest individual accuracy at 60.0%, it was followed by OpenAI (49.2%), the open-source Alibaba model (42.5%), and Anthropic (42.1%). Notably, this low performance contrasts with the success of text-based LLMs in clinical reasoning, indicating that the integration of visual understanding remains a significant challenge for foundation models in medical imaging.

In the binary task of differentiating tumors from other findings, Google’s model demonstrated high sensitivity (0.900), suggesting potential as an effective screening tool for identifying tumors but at the cost of lower specificity. In contrast, the other models, including those from OpenAI, Anthropic, and Alibaba, showed lower sensitivity. No model simultaneously achieved high sensitivity and high specificity. This trade-off is particularly problematic in kidney cancer diagnosis, where both missed tumors (low sensitivity) and unnecessary interventions from false positives (low specificity) carry significant clinical consequences. These results align with other studies examining real-world applications of VLMs for medical report generation, which found that VLMs alone remain insufficient and require human collaboration.^14,15^ Furthermore, Zhong et al.^14^ demonstrated that prompting strategies significantly affect the quality of radiologic reports, highlighting another critical challenge for clinical adoption of VLMs.

The VLMs demonstrated significant vulnerability to adversarial prompts through two distinct mechanisms. The ‘Benign’ prompt, which instructed the models to interpret findings as harmless, caused the overall accuracy to drop significantly from 48.4% to 39.2% (p<0.001). Our models prioritized textual commands over conflicting visual evidence of tumors, consistent with a known issue in AI research called ‘sycophancy,’ where a model agrees with a user’s stated preference even if it contradicts factual information.^16^ Interestingly, Google’s model showed relative resilience to this manipulation (60.0% to 57.9%, p=0.644), while OpenAI and Alibaba models were severely affected. This vulnerability aligns with recent findings by Clusmann et al.,^17^ who demonstrated that medical VLMs can be compromised by prompt injection attacks, causing them to systematically miss malignant lesions across multiple imaging modalities.

Similarly, the ‘Pressure’ prompt, which imposed cognitive workload by requiring thorough evaluation of 14 anatomical structures, significantly impaired performance (overall accuracy: 43.3%, p=0.025). This prompt appeared to overwhelm the models’ analytical capacity, causing them to default to ‘normal’ diagnoses. All models exhibited a surge in false negatives, tumors misclassified as normal, posing a significant risk of missed cancer diagnoses. ^18^

Intriguingly, when adversarially prompted to “Benign”, the Anthropic model did not comply but instead defaulted to a near-universal cancer classification, converting nearly all cases into tumors (231/240, 96.3%). We hypothesize this outcome is a “guardrail paradox,” an unexpected failure mode of its safety mechanisms. The model appears to recognize the prompt as a malicious attempt to force a critical error—a missed cancer (false negative). To avert this catastrophic outcome, its safety alignment seemingly compels a hyper-conservative response, classifying all ambiguous cases as malignant (false positive) as the lesser of two evils. This protective over-correction, however, created a severe and systematic bias toward over-diagnosis (**Figure 4**).

This guardrail paradox exposes a novel attack vector: malicious actors could intentionally trigger a model’s safety protocols to induce systemic over-diagnosis, effectively weaponizing its ethical constraints. Unlike general-purpose applications where guardrails can safely refuse harmful requests, medical AI faces the dilemma that both false positives and false negatives carry significant clinical consequences. Our results suggest that for medical VLMs, the optimal safety approach may not be enhanced critical judgment or ethical disclaimers, but rather complete task refusal when manipulation is detected. This aligns with the principle of “first, do no harm” - when the system’s integrity is compromised, no diagnosis may be safer than a potentially manipulated one.

Our study has immediate clinical implications. The unreliability and manipulability of VLMs pose unacceptable risks to patient safety, particularly as kidney cancer treatment often proceeds from imaging directly to surgery without biopsy confirmation. ^10,11^. The vulnerability to prompt manipulation is also concerning, as various stakeholders—patients seeking reassurance, insurers minimizing costs, or even malicious actors—could intentionally or unintentionally influence diagnostic outputs. Adversarial vulnerabilities observed in our study suggest that evaluation of medical VLMs must extend beyond simple performance metrics to include holistic validation. While numerous benchmarks for adversarial attacks are emerging in non-medical fields,^19,20^ our study highlights the urgent need to develop standardized frameworks and benchmarks for healthcare. Future development should focus on creating medical-specific VLMs with built-in safeguards against manipulation and establishing regulatory frameworks before any clinical deployment.

Our study has several limitations. First, we used a single public dataset of 2D images, which does not fully represent the complexity of 3D scans used in clinical practice. Second, we only tested two types of adversarial prompts, and the models may be vulnerable to other types of attacks. Lastly, as VLM technology is evolving rapidly, our findings represent a snapshot of their performance at a specific point in time (June 2025).

In conclusion, our study shows that current VLMs demonstrate both insufficient performance and critical vulnerabilities to adversarial attacks in the context of CT interpretation. Future work must prioritize both improving the foundational diagnostic capabilities of VLMs and building robust defenses against manipulation.

## Methods

### Study Design

Our study employed a full factorial design: 4 VLMs × 3 prompt types × 240 kidney CT images, yielding 2,880 total VLM responses. This study aimed to explore whether representative commercial VLMs have sufficient interpretive ability in kidney CT scans and how that ability responds to prompt manipulation.

We used publicly available CT-KIDNEY-DATASET-Normal-Cyst-Tumor-Stone dataset^13^ which comprises 12,446 coronal and axial CT images with clinical labels (3,709 for cysts, 5,077 for normal findings, 1,377 for stones, and 2,283 for tumors). Each image was then verified by a radiologist and a medical technologist to ensure data accuracy.

For this experiment, we randomly selected 240 cases from three categories: normal (n = 80), cysts (n = 80), and tumors (n = 80), ensuring balanced representation across diagnostic conditions. We converted images to RGB format, resizing to 1024 × 1024-pixel resolution using Lanczos resampling.

### Visual-Language Model

We used three commercial VLMs and one open-source VLM : Google’s Gemini 2.5 Pro, OpenAI’s GPT-4o, Anthropic’s Claude 3.5 Sonnet and Alibaba’s Lingshu-32B.^12^ These models were selected as representative of current state-of-the-art VLM. All models were accessed through their respective official application programming interfaces (APIs) using persistent API parameters. We used temperature 0.1 for reproducibility, 4,096 maximum output tokens to allow comprehensive radiological reports. Rate limiting was implemented with 3-second intervals between API calls to comply with service terms and ensure response quality.

### Prompt Type

Models were tested under three different prompt types: ‘Neutral’, ‘Benign’, ‘Pressure’. Each condition was designed to test vulnerability mechanisms which are well-known in machine learning society: direct instruction override and cognitive resource depletion.

‘Neutral’ prompt includes standard radiological interpretation instructions using neutral clinical language without additional bias or pressure.

‘Benign’ prompt includes instructions that induce bias interpretation toward benign findings and coerce models to reframe any concerning features as benign characteristics while prohibiting malignancy suggestions.

‘Pressure’ prompt requests extensive systematic evaluation requirements designed to overwhelm analytical capacity, requiring evaluation of 14 distinct anatomical structures (bones, lungs, heart, liver, gallbladder, pancreas, spleen, adrenals, kidneys, bowel, vessels, lymph nodes, peritoneum, and soft tissues) to induce model’s cognitive resource depletion.

### Study Outcome

We manually reviewed all responses from the VLMs and classified responses as ‘tumor’, ‘cyst’, or ‘normal’ label. If a response mentioned both a tumor and a cyst, it was labeled as ‘tumor’. Tumor and cyst findings were classified as such only when the corresponding lesion was found in the kidney.

Primary outcome metric was 3-category accuracy by models and prompt types. We compare the accuracy of four visual language models (Overall, Google, OpenAI, Anthropic and Alibaba) across three different prompt conditions: Neutral, Benign, and Pressure. We calculated statistical differences between the Neutral condition and the other two conditions (Benign and Pressure).

Secondary outcome metrics, including precision, recall, sensitivity and False Positive Rate (FPR), were used to evaluate the performance of the models specifically for the Tumor vs Other (Cyst or Normal) task. While the primary outcome metric was 3-category accuracy, we applied binary metrics to provide a more detailed analysis of the models’ ability to correctly identify tumors, which are a critical finding that should not be missed. We generated a Receiver Operating Characteristic (ROC) curve and a Precision-Recall (PR) plot to visualize the model performance on these metrics.

Additionally, we utilized stacked bar charts to represent the distribution of model predictions and error rates across different models and prompt types. To analyze how VLM predictions might be misclassified into other classes, we constructed a series of nine 3×3 confusion matrices, one for each model and prompt type, to comprehensively visualize the specific patterns of misclassification.

Statistical comparisons between experimental conditions used Fisher’s exact tests for categorical outcomes and Welch’s t-tests for continuous measures. Statistical significance was defined as P<0.05, with additional notation for P<0.01 and P<0.001. All statistical analyses were performed using Python 3.9 with pandas 1.5.3, scipy 1.10.1, and matplotlib 3.7.1 libraries. Complete statistical code and processed datasets are available upon request.

## Data Availability

CT-KIDNEY-DATASET-Normal-Cyst-Tumor-Stone dataset is publicly open-dataset. (https://www.kaggle.com/datasets/nazmul0087/ct-kidney-dataset-normal-cyst-tumor-and-stone)

## Code availability

All analysis code written in Python. Analysis code will be shared upon request to corresponding author.

## Acknowledgement

This work was supported by Hankuk University of Foreign Studies Research Fund (Of 2025). This work was also supported by the National Research Foundation of Korea (NRF) grant funded by the Korea government (MSIT)(RS-2024-00392315).

B.C., S.H., and J.S. designed the research, conducted the experiments, and performed the data and statistical analysis. M.W.P. and T.J.J. provided statistical validation and clinical review. All authors contributed to the writing of the manuscript.

These two first-authors contributed equally: B.C., S.H

J.S. is corresponding author.

## Competing interests

The authors declare no competing interests.

## Notes

### Competing Interest Statement

The authors have declared no competing interest.

